# Persistent Aerointolerant Gut Microbes and Butyrate Depletion in Severe Acute Malnutrition

**DOI:** 10.1101/2025.02.07.25321872

**Authors:** Aminata Camara, Salimata Konate, Maryam Tidjani Alou, Nicholas Armstrong, Aurelia Caputo, Amadou Hamidou Togo, Safiatou Niare, Abdoulaye Kassoum Kone, Issa Traore, Anthony Levasseur, Ogobara K. Doumbo, Mahamadou Ali Thera, Matthieu Million

**Author notes:** **Corresponding author:**; IHU - Méditerranée Infection ME-PHI, Aix-Marseille University, 19-21 Boulevard Jean Moulin, 13005 Marseille.

## Abstract

Severe acute malnutrition (SAM) constitutes the leading risk for childhood mortality worldwide. Butyrate, a critical metabolite for mucosal immunity produced by anaerobic gut microbes, has been found to be dramatically depleted in children experiencing fatal outcomes related to SAM. However, the association between butyrate depletion and gut microbiota alteration has not yet been fully explored. Here, we performed a large group-matched case-control study in children aged 0-59 months in Mali. We compared faecal short-chain fatty acids (SCFAs) and gut microbiota from children with SAM and healthy controls, using gas chromatography-mass spectrometry and 16S rRNA v3v4 gene amplicon sequencing. A total of 253 children, including 143 with SAM and 110 controls, were included. Our findings revealed a dramatic imbalance in gut microbiota characterised by the predominance of aerotolerant micro-organisms in children with SAM, irrespective of age and sex. Importantly, aerointolerant species were totally absent in eight samples from children with SAM. A pronounced depletion of faecal valeric, butyric, propanoic and, to a lesser extent, acetic acid was observed, correlating with a ten-fold decrease in butyrate-producing microbes among the SAM population. This notable depletion of gut aerointolerant prokaryotes and butyrate levels persisted without being reversed at the time of discharge. Future therapeutic strategies aimed at addressing SAM should focus on restoring the gut anaerobic microbiota and faecal propionate and butyrate.

## 1. Introduction

Severe acute malnutrition (SAM) is a neglected tropical disease around the world, with more than 50 million acute malnutrition cases annually [1]. SAM represents the leading risk factor for attributable disability-adjusted life-years and childhood mortality globally [2]. This condition is defined by a weight-for-height measurement below three standard deviations from the World Health Organization (WHO) standards and/or a mid-upper arm circumference (MUAC) of less than 115 mm and/or the presence of bilateral oedema [3]. Decades of research have identified inadequate breastfeeding and diet as predisposing factors contributing to SAM [4]. Beyond the lack of macronutrients which lead to protein-energy malnutrition, SAM has been associated with oxidative stress, the accumulation of free radicals, and mitochondrial damage in the liver, particularly in its more severe form, kwashiorkor [5]. These findings prompted the addition of several micronutrients exhibiting direct or indirect antioxidant properties into ready-to-use therapeutic food (RTUF) [3]. Notwithstanding improvements in antibiotic and nutritional management, SAM remains a fatal disease with a substantial number of cases which are refractory to recommended therapy [6].

We conducted a review focusing on the specific features of marasmus and kwashiorkor, the two clinical manifestation of SAM [7]. Through a comparative meta-analysis, we found that kwashiorkor and marasmic-kwashiorkor, the most severe forms of SAM with bilateral oedema, were associated with fatty liver. Fatty liver in malnutrition has been linked to peroxisomal and mitochondrial dysfunction [8]. However, the underlying mechanisms linking inadequate diet and liver peroxisomal and mitochondrial dysfunctions remain unelucidated. It is essential to highlight the neglected role of gut microbiota in this context.

Butyrate has been identified as a key microbial metabolite with several beneficial effects for mammal health, notably for mucosal immunity [9], liver mitochondrial integrity, and liver fat regulation [10,11]. The protective role of butyrate in SAM is underscored by research indicating a link between faecal butyrate depletion and mortality among children with this condition [12]. Butyrate is mainly produced by gut aerointolerant [13] prokaryotes [14].

Preliminary studies using 16S rRNA gene v3v4 amplicon sequencing and culturomics deciphered a dramatic aerointolerant depletion in SAM [15,16]. Thus, we hypothesised that gut aerointolerant depletion could be associated with faecal butyrate depletion in SAM. However, to our knowledge, no prior studies have assessed faecal SCFAs comparing children with SAM and healthy controls, nor have any studies linked gut aerointolerant prokaryotes and faecal short-chain fatty acids in SAM. Accordingly, we performed a large case-control study of children with SAM, comparing their faecal microbial content using 16S amplicon sequencing, and their faecal short-chain fatty acid content using gas chromatography-mass spectrometry.

## 2. Materials and methods

### 2.1. Participants and study design

The design, setting, and location of this case-control MALISAM (for Mali and SAM) study has been previously reported [17] in a study focusing on *Methanobrevibacter smithii*. The present study focuses on aerointolerant and butyrate-producing microbes and short-chain fatty acids. This case-control study was reported according to the recommendations in the STROBE Statement [18], between May and August 2015, and then between August 2016 and January 2017, in a peri-urban area of Kalaban Coro located in the southeast of Bamako in Mali. Participants with moderate acute malnutrition, and those who refused to give consent, or withdrew were excluded. Children were matched by sex and age ranges of 0-6 months, 7-12 months, 13-24 months and >24 months.

### 2.2. Ethical considerations

Ethics Committee of the University of Bamako Faculty of Medicine and Odonto-Stomatology waived ethical approval for this work on 22 May 2014 (2014/46/CE/FMPOS). Informed and signed consent was obtained from all legal representatives (parents).

### 2.3. Data sources, measurement and definitions

SAM cases and controls were defined following WHO guidelines, as described in our previous study [17]. Anthropometric parameters including weight, height, mid-upper arm circumference, and age were used to determine nutritional phenotypes based on the WHO Anthro software which calculates the ratios of weight/age, weight/height and height/age according to the date of inclusion, gender, date of birth and the presence or not of oedema. All the children were screened over the same period and in the same geographical area in order to address potential sources of bias.

### 2.4. Gut microbiota characterisation

#### 2.4.1. V3v4 amplicon sequencing

Total DNA was extracted from the stool samples using two protocols [19]. The first was based on a physical lysis with glass powder followed by an overnight chemical lysis with proteinase K. The resulting solution was washed and eluted according to the recommendations of the manufacturers of the Macherey-Nagel DNA Tissue extraction kit [19]. For the second protocol, glycoprotein lysis and deglycosylation steps were added to the first protocol [19]. Sequencing of the resulting DNA from these two protocols was performed, targeting the V3-V4 regions of the 16S rRNA gene, as previously described [15]. All reads from the two protocols were grouped and clustered with a threshold of 97% identity to obtain operational taxonomic units (OTU). All OTUs consisting of <20 reads were removed. The remaining OTUs were blasted against SILVA123 and the Meta GX culturomics database and assigned to a species if they matched one with at least 97% identity.

#### 2.4.2. MetaGX pipeline

MetaGX is a home-made software pipeline developed with the start-up XeGen (http://xe-gen.eu/), based on QIIME [20], using BLAST [21] for taxonomic assignment and SILVA [22,23] as a reference dictionary. To have a single method of data analysis for v3v4 amplicon sequencing of 16S RNA gene, this pipeline provides a large database for inter-microbial comparisons or unsupervised analyses between bacterial species. Compared to the usual version of QIIME, the pipeline has been adapted to include a dictionary of species discovered in our laboratories using the culturomics method, which is updated continuously. In addition, the pipeline has been specifically designed to name all OTUs, including those that do not correspond to a cultivated species (referred to as IHU_PS_ XX_YY, where “IHU” stands for Institut Hospitalo-Universtaire (our institute), “PS” stands for putative species, “XX” is the percentage of similarity of the OTU with the closest cultivated species, with their most precise taxonomy, followed with “YY”, an arbitrary number (such as 1, 2, 3, etc.). For example, IHU_PS_96_Lactobacillus23876). This makes it possible to potentially discover OTUs which are important in health or disease even if no accurate taxonomic assignment is possible. The version used here (the second version of the pipeline and second version of the dictionary, V2D2) included 556 new culturomics species. This pipeline has already been used in a recent article on the comparison of culturomics and metagenomics [24]. This second version of the pipeline introduces the concept of “multi-assignment”. When using BLAST and the best BLAST hit, the selection of the best BLAST hit can lead to aberrant assignments (*Shigella sonnei* instead of *Escherichia coli,* due to uncertain bases in the reference sequence of *Escherichia coli* in SILVA). Knowing that the hypervariable v3v4 region of the 16S ribosomal RNA gene (like any region and even the 16S rRNA gene as a whole) sometimes cannot discriminate between several species, we favoured an assignment at the species level rather than limiting assignment to the lowest rank. To do this, the software was set up to return all known species with less than 0.5% divergence from the BBH (best BLAST hit), with respect to the 98% similarity criterion.

#### 2.4.3. Assessment of aerointolerant/aerotolerant distribution and imbalance

We used a previously published database from our laboratory [15] to classify the OTUs detected in our samples as either aerointolerant or aerotolerant. This database has been updated and is available online (https://www.mediterranee-infection.com/acces-ressources/base-de-donnees/list-of-prokaryotes-according-to-their-aerotolerant-or-obligate-anaerobic-metabolism). For species that did not exist in this database, we consulted online databases such as LPSN (list of prokaryotic names with standing in nomenclature, https://www.bacterio.net, last accessed February 7^th^, 2025) and BacDive (bacterial phenotypic data for high-throughput biodiversity analysis, https://bacdive.dsmz.de, last accessed February 7^th^, 2025) to complete the classification. For OTUs taxonomically assigned below the species level, putative species belonging to a cluster with cultured members having all the same oxygen tolerance were assigned to this phenotype.

To assess the aerointolerant/aerotolerant distribution and imbalance in large scale sequencing datasets with assignment at the species level, we previously proposed the calculation of two markers [15,25]. The first, was at the individual level, where we proposed the metagenomic aerotolerant predominance index (MAPI) which was the ratio between the relative abundance of aerotolerant prokaryotes and the relative abundance of aerointolerant prokaryotes [25]. The MAPI has been used by other teams and has helped to decipher an aerointolerant depletion in Ugandan children with language development impairment [26]. As we observed few children with SAM, but no healthy controls, without any sequences corresponding to aerointolerant species, we proposed a modified index, the metagenomic aerointolerant predominance index (MAIPI, Equation (1)). The logarithmic transformation made it possible to obtain a normal distribution in controls, as previously reported [25].

**Metagenomic aerointolerant predominance index (MAIPI)**

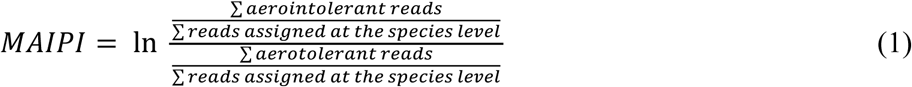

For samples with zero aerointolerant reads, the MAIPI was calculated as follows (Equation (2)), and this was noted in tables and figures.

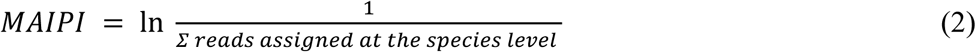

Secondly, we proposed the aerotolerance odds ratio, (Table 1, Equation (3)) at the disease level, by comparing the proportion of aerotolerant and aerointolerant prokaryotes among species enriched or depleted in malnourished children or in healthy controls for any disease associated with dysbiosis [25]. In contrast, the AOR characterises dysbiosis associated with a disease and not an individual. In our previous works on SAM [15,25], the study of these two markers was consistent in the direction of depletion of aerointolerant bacteria, but the sample size of the present study is much larger and controls for confounding factors much more effectively.

**Table 1.** Aerotolerant Odds Ratio (AOR)

| Species | Enriched in<br>disease<sup>1</sup> | Depleted in<br>disease<sup>1</sup> |
| --- | --- | --- |
| Aerotolerant | A | B |
| Aerointolerant | C | D |

<sup>1</sup> Determined here by frequency difference (Fisher exact or chi-square test according to sample size). A = number of aerotolerant species enriched in the disease, etc...

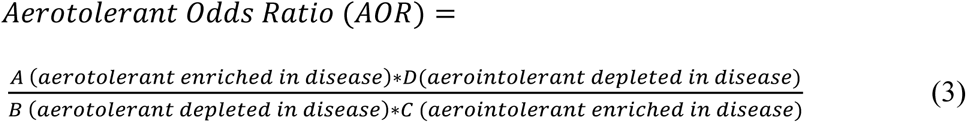

An AOR≠1 with a *P*-value < .05 suggests a significant aerointolerant/aerotolerant imbalance. An AOR > 1 is synonymous with an aerotolerant relative enrichment in the disease. Diseases associated with oxidative stress are expected to have an AOR > 1.

### 2.5. Short-chain fatty acid assessment by gas chromatography-mass spectrometry

The acetic, propanoic, butyric, isobutyric, valeric, isovaleric, caproic, and heptanoic quantities of the short-chain fatty acids (SCFAs) were assayed in the stool samples using gas chromatography coupled with mass spectrometry (GC Clarus 500, SQ 8 S mass spectrometer, Perkin Elmer, Villebon-sur-Yvette, France), as previously reported [27]. Briefly, we used a quantification method by internal calibration which consisted of preparing six range points (0.1 MM-10 mM) and two controls to which an internal standard (2-ethylbutanoic acid) was added. Concerning the samples, 500 mg of stool was diluted in 2.5 mL of high pressure liquid chromatography (HPLC) water and then suspended before adjusting the pH to between two and three with 37% hydrochloric acid. After centrifugation at 5000 rpm for 20 minutes, 1.5 mL of supernatant was collected and then centrifuged again at 13 000 rpm for five minutes. Finally, 1 mL of clear and limpid supernatant was recovered and doped with the internal standard before being placed in GC vials for analysis. Turbid samples (those containing debris after centrifugation) were passed through 0.2 μm filters. Compounds were then separated on an Elite-FFAP column (30 m, 0.25 mm ID, 0.25 mm film thickness) using helium as a carrier gas. SCFAs were selectively monitored by selected ion recording mass spectrometry. All data was collected and processed using Turbomass 6.1 (Perkin Elmer).

### 2.6. Statistical methods

Visual examination of distributions and the Shapiro-Wilk normality test were used for the distribution of quantitative data to apply parametric tests (t-test) or nonparametric tests (Mann-Whitney test). The chi-square test was used to test the differences in proportion between the groups. Linear regression was used to test associations between age and each SCFA. Partial least square regression (PLS-R) and the random forest model were used to investigate the association between SCFA and age in controls. A *P*-value ≤ .05 was considered significant. Statistical analyses were performed using SPSS software version 20.0 (IBM, Paris, France), GraphPad Prism 9.0 (GraphPad Software, La Jolla, USA) and XLSTAT 2022.3.1 (Addinsoft, Paris, France).

## 3. Results

### 3.1. Participants and general results

Of a total of 180 malnourished and 209 healthy children were screened and 143 severe acute malnourished and 110 healthy infants were finally included for biological analysis. A total of 253 children were therefore ultimately included. Demographic data are available in reference [17]. It was possible to analyse a second sample for the gut microbiota profile at the time of discharge for 47 children (41 SAM cases and 6 controls), and v3v4 16S rRNA sequencing was thus performed on 300 samples yielding 17 123 617 high-quality filtered reads (14 453 582 from the first samples and 2 670 035 from the second samples). SCFA analysis was performed for the 110 controls but only for 140 malnourished children due to an insufficient amount of stool samples. A second sample was analysed for SCFA at the time of discharge for 39 children (34 SAM cases and 5 controls). Accordingly, 279 samples were analysed for SCFA profiling.

### 3.2. Gut dysbiosis is characterised by an aerointolerant/aerotolerant imbalance, a relative enrichment in potential pathogens and a depletion of potential probiotics in SAM

#### 3.2.1. Depletion of gut aerointolerant prokaryotes in SAM

Using v3v4 16S rRNA amplicon sequencing of the ribosomal 16S RNA data, we found that total number of reads was not different between cases and controls (median, 46 757 for cases and 45 890 for controls). However, we confirmed the dramatic depletion of the relative abundance of aerointolerant species in severely malnourished children (Figure 1). Comparing the relative abundance of gut microbes according to aerotolerance, we found that the relative abundance of aerointolerant gut microbes was six-fold higher in controls (median, 63.0%, interquartile range [IQR 46.5%-77.3%] vs 11.6% [2.4%-41.5%], Mann-Whitney test, *P* < .0001), and two-to three fold lower for aerotolerant ones (34.5% [18.7%-51.2%] vs 86.8% [56.4%-96.2%]).The relative abundance of OTUs for which aerotolerance could not be determined was slightly but not significantly higher in controls (1.0% vs 0.7%).

**Figure 1.**
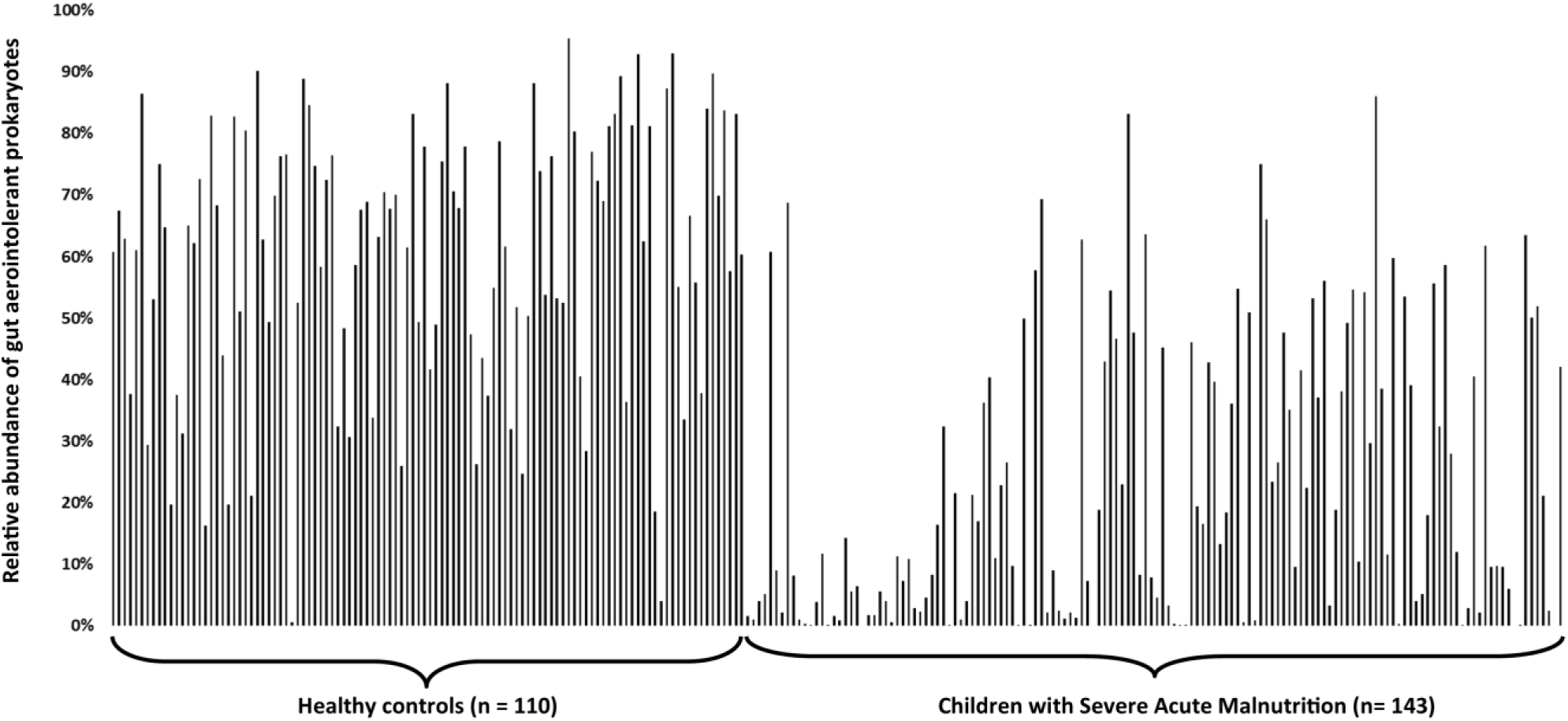
Relative abundance of gut aerointolerant prokaryotes in children with SAM and healthy controls.

#### 3.2.2. Aerointolerant/aerotolerant imbalance at the individual level

We then assessed the metagenomic aerointolerant predominance index (MAIPI). Healthy children had a large and significant predominance of aerointolerant bacteria (MAIPI, median [interquartile range], 0.63 [-0.08 to 1.47], one sample Wilcoxon test for MAIPI ≠ 0, *P* < .0001). Children with SAM had a very large and significant predominance of aerotolerant bacteria (−2.02 [-3.70 to -0.32], *P* < .0001, Figure 2a). Surprisingly, age was not associated with MAIPI neither in SAM nor in healthy children as assessed by univariate linear regression. Healthy children had an aerointolerant predominance as early as one month of age (Figure 2b).

**Figure 2:**
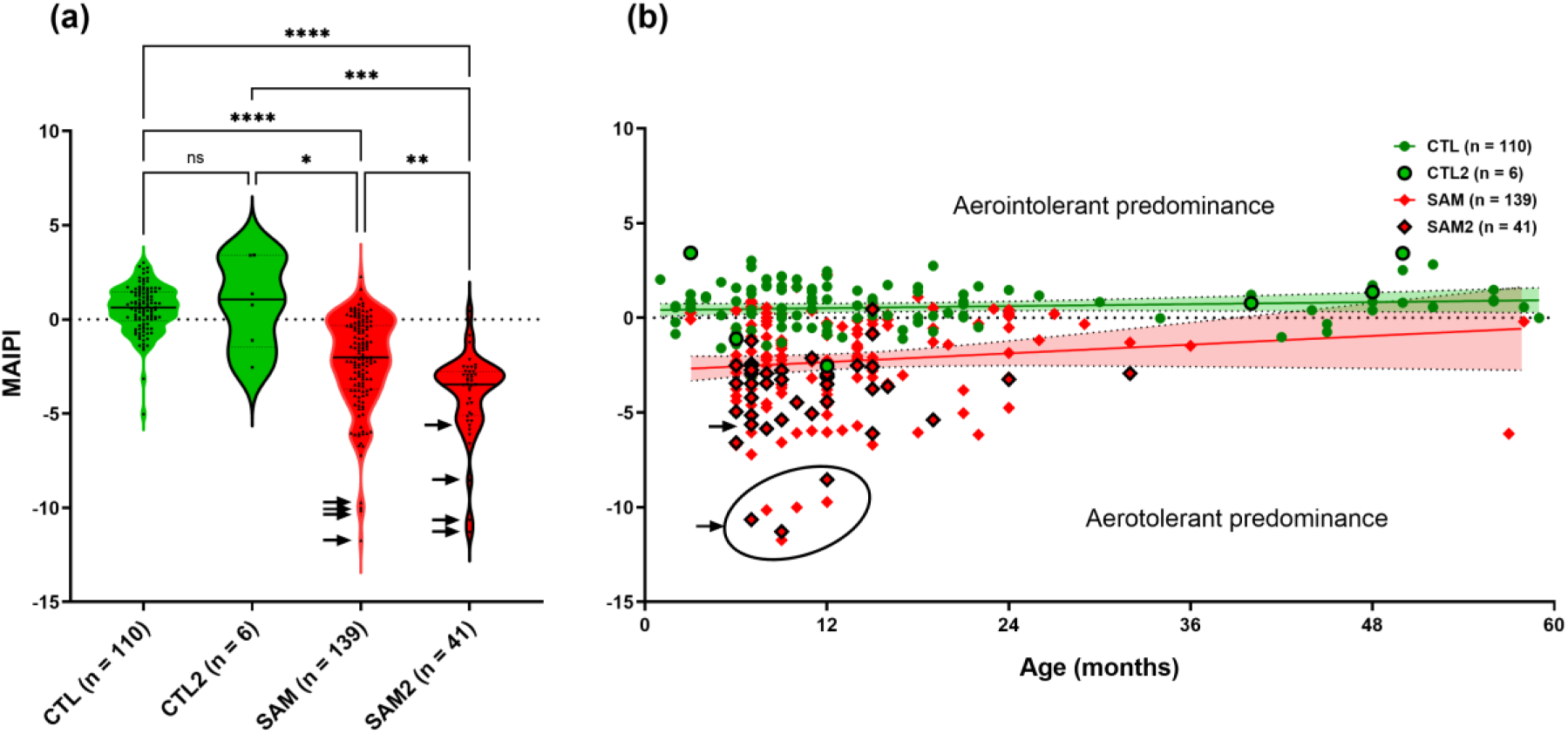
Metagenomic aerointolerant predominance index (MAIPI) in healthy controls versus children with severe acute malnutrition. MAIPI = ln (relative abundance of aerointolerant species/relative abundance of aerotolerant species). CTL: healthy controls, CTL2: second sample collected for six control children, SAM: Severe acute malnutrition, SAM2: second sample collected at the time of discharge in 41 children with SAM. **Arrows**: samples without any v3v4 16S rRNA reads assigned to any aerointolerant species (the MAIPI was calculated as if an additional read would have been aerointolerant, see Methods). **(a)** Univariate comparison. Two-tailed Mann-Whitney test. Median and interquartile range are shown. \**P* < 0.05, \*\**P* < 0.005, \*\*\**P* < 0.0005, \*\*\*\**P* < .0001. **(b)** Linear regression according to age. Age was not significantly associated with MAIPI either in healthy controls (coef. 0.0075, *P* = 0.24) or in children with SAM (coef. 0.022, p = 0.13).

Thanks to the MAIPI, cases and controls were using univariate and multivariate models. In the univariate analysis, the MAIPI of healthy controls was significantly higher than the MAIPI of children with SAM (Mann-Whitney test, *P* < 10^-3^, Figure 2a). This was confirmed in a multivariate linear regression model including age, sex, and nutritional status as potential predictors and the aerointolerant/aerotolerant balance as an outcome (MAIPI). Only nutritional status (SAM) and sex but not age were identified as independent predictors of MAIPI (SAM, coefficient: -2.72, 95% confidence interval (95%CI) -2.21 to - 3.24, *P* = 3.0 x 10^-21^). This definitively confirms that SAM is associated with a severe aerotolerant/aerointolerant imbalance with an aerotolerant relative enrichment. When assessing the MAIPI in the second samples collected at the time of discharge in 47 children (41 children with SAM and 6 healthy controls), we observed that treatment did not correct the aerointolerant/aerotolerant imbalance in these children (Figure 2).

#### 3.2.3. Most species depleted in SAM are aerointolerant

Of the 290 species or putative species for whom frequency was significantly different between SAM and controls, aerotolerance could not be determined for 52 of them. Of the 238 species for whom aerotolerance could be determined, gut aerointolerant microbes were much more likely than aerotolerant ones to be depleted in SAM (Table 2, Aerotolerant Odds Ratio = 28.7, 95% CI (13.9 – 63.4), *P* < .10^-7^). Comparing species enriched in controls or children with SAM, 70.7% of the species enriched in controls were aerointolerant, while only 22.5% of species enriched in children with SAM were aerointolerant.

**Table 2.** Number of species enriched or depleted in SAM according to aerotolerance among 238 species with a significant different frequency in healthy children and children with SAM

| Species | Enriched in SAM<sup>1</sup> | Depleted in SAM<sup>1</sup> |
| --- | --- | --- |
|  | (n = 89) | (n = 149) |
| Aerotolerant | 78 (87.6%<sup>2</sup>) | 29 (20.9%<sup>3</sup>) |
| Aerointolerant | 11 (12.4%<sup>2</sup>) | 120 (79.1%<sup>3</sup>) |

SAM: Severe acute malnutrition. <sup>1</sup>determined by frequency difference (Two-tailed Fisher exact or chi-square test according to sample size). <sup>2</sup>Of the 89 species enriched in SAM. <sup>3</sup>Of the 149 species depleted in SAM.

#### 3.2.4. Absence of aerointolerant gut microbes in eight samples from children with SAM

Strikingly, eight samples from children with SAM (but none from healthy children) had no reads associated with aerointolerant species (Table 3). As indicated by the number of total assigned reads, this was not associated with a low efficiency of 16S amplification. Focusing on the most abundant species in these samples, we found that the most represented OTUs were assigned to *Escherichia coli* and *Streptococcus salivarius*. In two children a single species had a relative abundance > 90% ([*E. coli*, *S. dysenteriae*, *S. flexnerii*] 97.9%, [*Pseudomonas lactis*] 93.9%). In one child, *Morganella morganii*, a Pseudomonadota known to resist amoxicillin (AmpC β-Lactamase), was the majority species with > 75% relative abundance (Table 3).

**Table 3.** Characteristics of the eight samples without any reads assigned to an aerointolerant species

| Participants<sup>1</sup> | Total reads | Species corresponding to the most abundant OTUs<sup>2</sup> |
| --- | --- | --- |
| SAM_035 (8,M) | 25705 | [*E. coli*, *Shigella dysenteriae*, *Shigella flexnerii*] 38%<br>[*Pseudomonas pseudoalcaligenes*] 27%<br>[*Streptococcus salivarius*] 26% |
| SAM_039 (9,F) | 125697 | [*E. coli*, *S. dysenteriae*, *S. flexnerii*] 97.9% |
| SAM_052 (12,F) | 16861 | [*E. coli*, *S. dysenteriae*, *S. flexnerii*] 55.7%<br>[*S. salivarius*] 31% |
| SAM_061 (10,F) | 22438 | [*S. salivarius*] 45.8%<br>[*Pediococcus acidilactici*, *Pediococcus pentosaceus*] 28.7%<br>[*E. coli*, *S. dysenteriae*, *S. flexnerii*] 11.6% |
| SAM_016 (7, F) | 42235 | [*S. salivarius*] 60.3%<br>[*Bifidobacterium longum*] 18.3%<br>[*E. coli*, *S. dysenteriae*, *S. flexnerii*] 13.9% |
| SAM_085 (9,M) | 80323 | [*Pseudomonas lactis*] 93.9% |
| SAM_086 (7,M) | 273 | [*Halomonas pacifica*] 43.6%<br>[*E. coli*, *S. dysenteriae*, *S. flexnerii*] 41%<br>[*Shewanella indica*] 15.4% |
| SAM_093 (12,M) | 5128 | [*Morganella morganii*] 76.3% |

<sup>1</sup>Age (months), sex. <sup>2</sup>Operational taxonomy units (OTUs) with a relative abundance > 10%. For each OTU, the most probable species (known to be associated with humans) are shown.

#### 3.2.5. Species enriched in SAM

The top three OTUs enriched in SAM matched well-known potentially pathogenic enterobacteria. *Klebsiella pneumoniae* was repeatedly identified as a possible taxonomic assignment for all three top OTUs (Figure 3). The most enriched OTU may also assigned to *Citrobacter koseri, Shigella flexneri, Shigella sonnei*, and *Serratia marcescens*. The second OTU may also correspond to *Klebsiella oxytoca*. The third most enriched OTU may also correspond to *Salmonella enterica*. In addition to enterobacteria, other pathogens have been found in species associated with malnutrition. These species included *Fusobacterium nucleatum* from the phylum of Fusobacteria. This species is increasingly associated with dysbiosis, infection and cancer [28]. Finally, we found enrichment of the microbiota of malnourished children with *Corynebacterium kroppenstedtii*, a bacterium associated with female breast infections and maternal dysbiosis [29,30].

**Figure 3:**
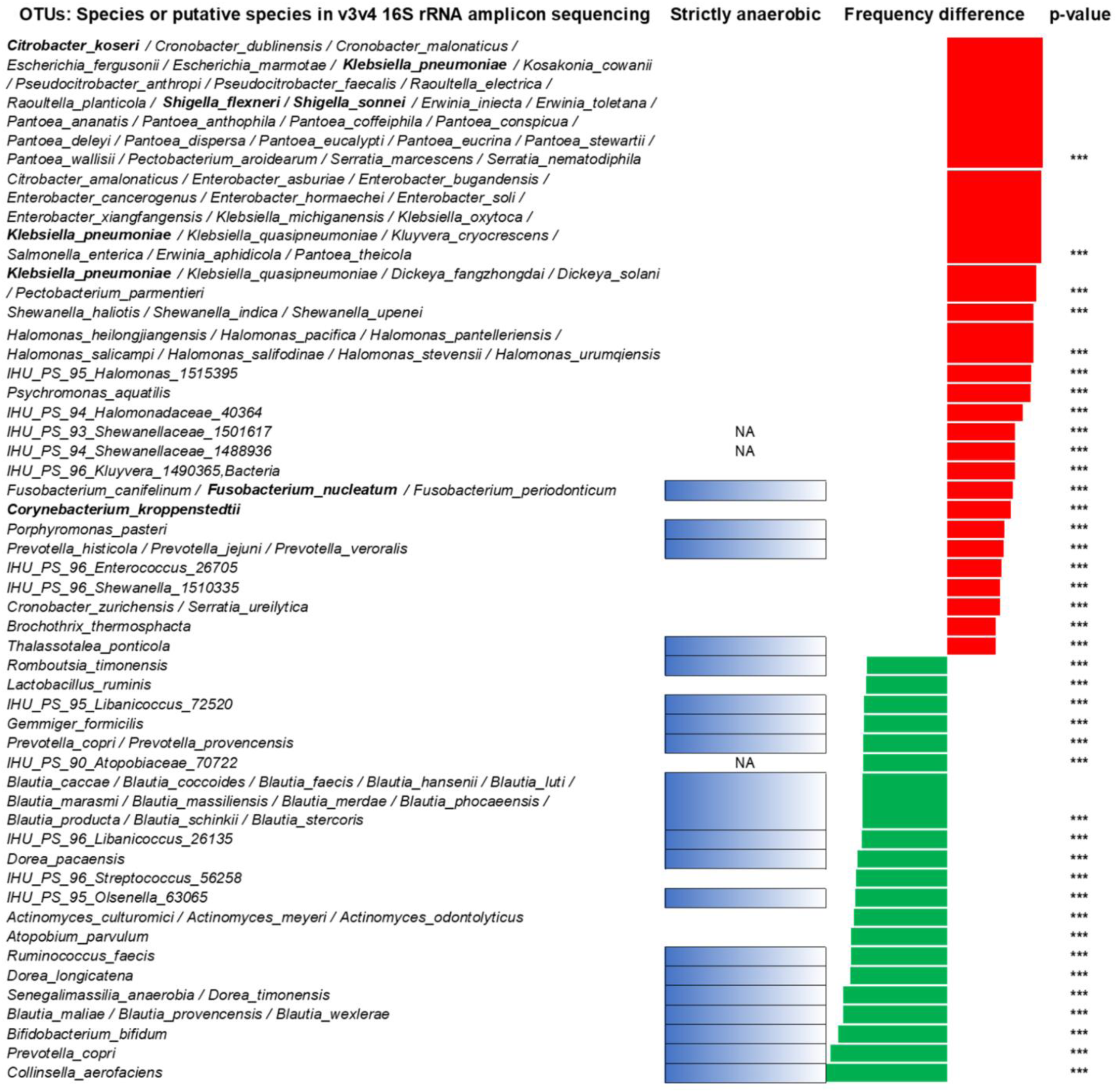
Frequency difference between species detected in infants with and without SAM. Only the first 20 species significantly associated with each group are shown. In green, species or putative species significantly depleted in SAM, in red, species or putative species significantly enriched in SAM. \*\*\**P* < .0005. Bold: species associated with disease (pathogenic species).

#### 3.2.6. Species enriched in healthy controls

At the species level (Figure 3), we analysed the top 20 species enriched in healthy controls. We found that aerointolerant species/genus isolated in our centre using the microbial culturomics approach were enriched only in healthy controls, including the highly abundant *Romboutsia timonensis* [31], *Dorea pacaensis*, and *Senegalimassilia anaerobia* [32]. The potential probiotics *Bifidobacterium bifidum* and *Ligilactobacillus ruminis* were also enriched in controls. *Faecalibacterium prausnitzii*, the typical human aerointolerant symbiont [33], acetate consumer and butyrate producer, was strongly depleted in SAM. We identified two members of the *Christensenellaceae* family (IHU_PS_91_Christensenellaceae_2234 and IHU_PS_92_Christensenellaceae_2858) enriched in controls. This family corresponds to inherited microbes associated with normal body mass index and adiposity in obesity studies [34]. Consistently with our previous findings [15,17], the aerointolerant archaeal *Methanosphaera stadtmanae* was also enriched in controls. Strikingly, no *Pseudomonadota* were found enriched in controls.

### 3.3. Faecal propionate and butyrate (but not acetate) increases with age in healthy children but not in children with SAM

#### 3.3.1. Short-chain fatty acids associated with age in healthy children

First, we evaluated the evolution of the faecal concentration of the different SCFAs with age in healthy children. Using a PLS-R and a random forest model, only propionate and butyrate, but not acetate and the other SCFAs, were predictors of child age (Figure 4).

**Figure 4:**
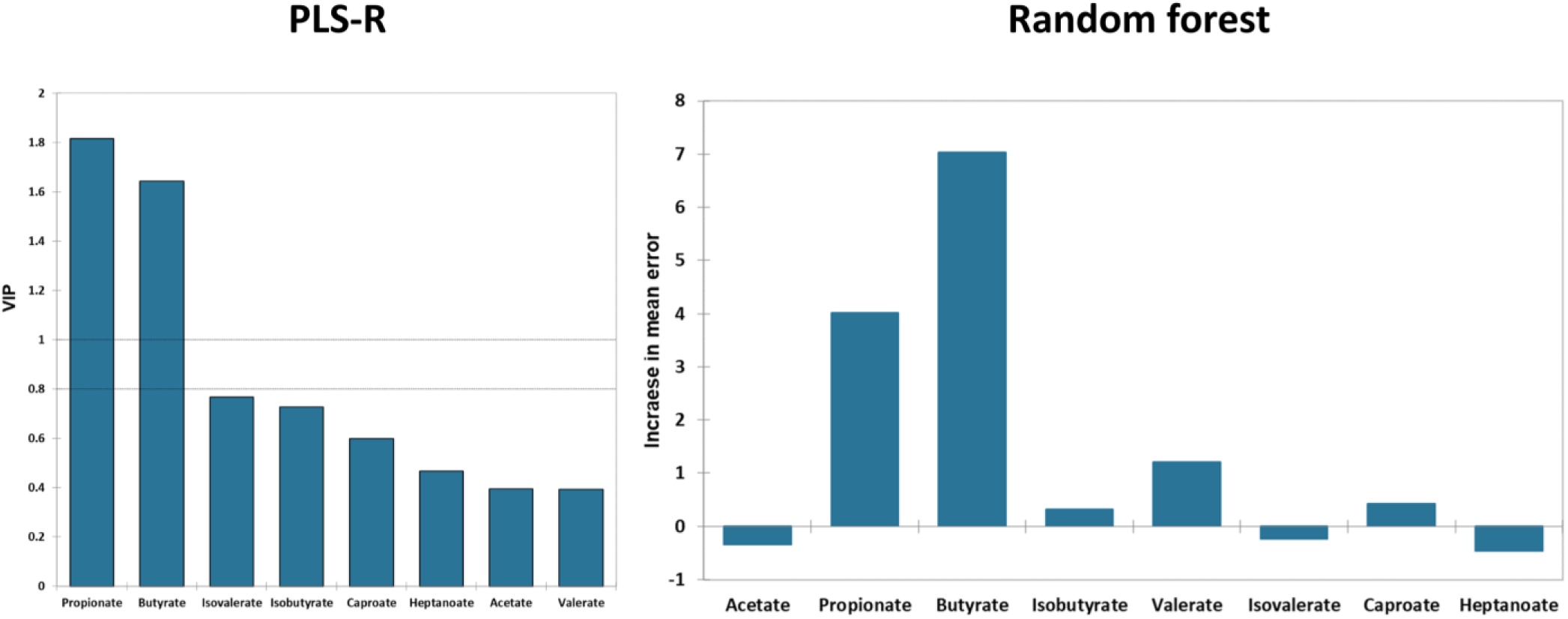
Association between each faecal SCFA and age in healthy controls. Left panel: PLS-R: partial least square regression, VIP: variable importance for prediction. Right panel: random forest model. Concentration of each SCFA in mM. Propionate and butyrate, but not acetate, are the most important variables for age.

#### 3.3.2. Species correlated with faecal butyrate

We evaluated which species correlated with faecal butyrate concentration. *Collinsella aerofaciens* and *Faecalicatena intestinalis* were the two species with the highest correlation coefficient. Conversely, *Vibrio litoralis* was one of the species detected by sequencing with the largest negative correlation coefficient.

#### 3.3.3. Short-chain fatty acids and severe acute malnutrition

All short-chain fatty acids were depleted in malnourished children in comparison to control children (Figure 5). The decrease of valeric acid (−100%), butyric acid (−88%) and propanoic acid (−85%) was more dramatic than for acetic acid (−58%). Faecal propionate (*P* < .001) and butyrate (*P* = .0002) increased with age in controls but not in children with SAM (Figure 6). The slope of increase of butyric acid was six times higher in controls than in malnourished children (slope of the linear regression, 0.06 vs 0.01) with a significant difference between the two groups, *P* = .05.

**Figure 5:**
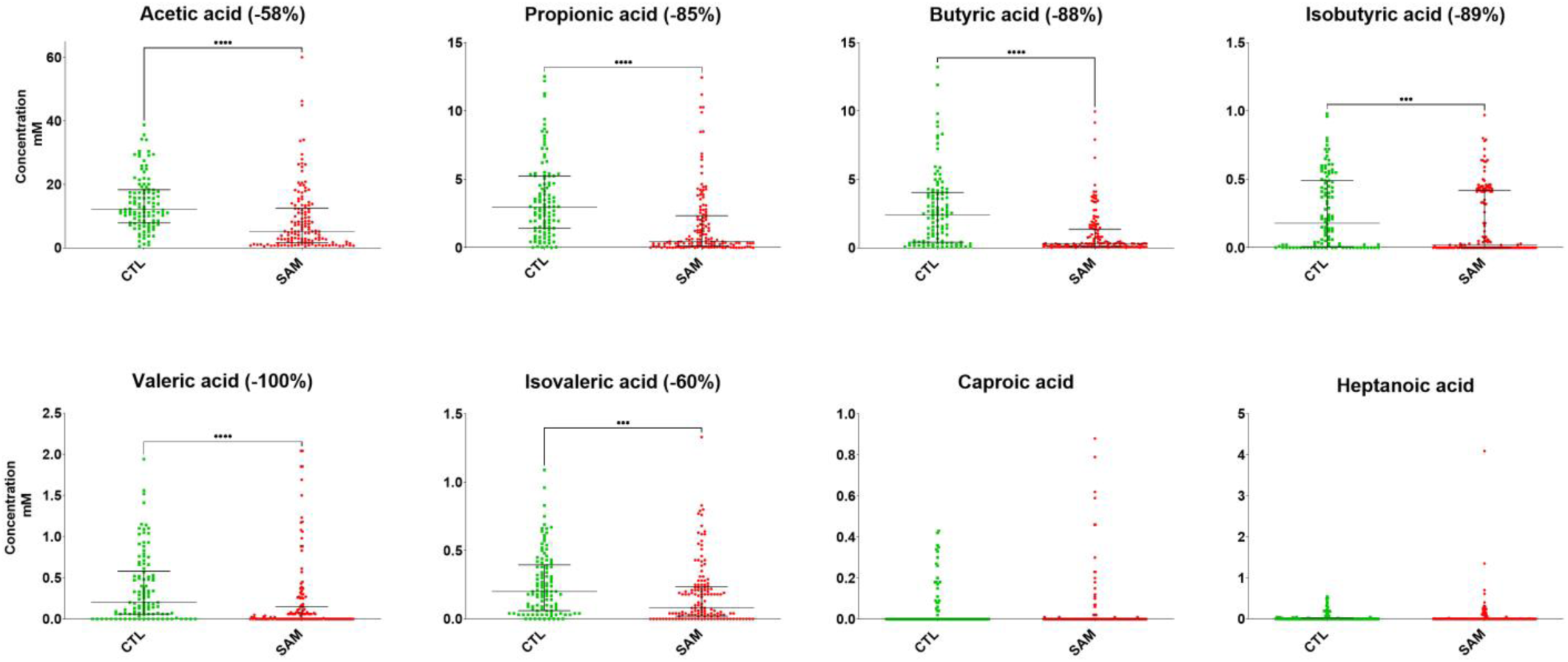
Short-chain fatty acid concentration in stool samples of healthy controls and children with severe acute malnutrition. mM: millimolar, CTL: healthy controls (n = 110), SAM: children with SAM (n = 140). Median and interquartile ranges are shown; Mann-Whitney U test. \*\*\**P* < .001, \*\*\*\**P* < .0001. There was no significant difference for caproic and heptanoic acid.

**Figure 6.**
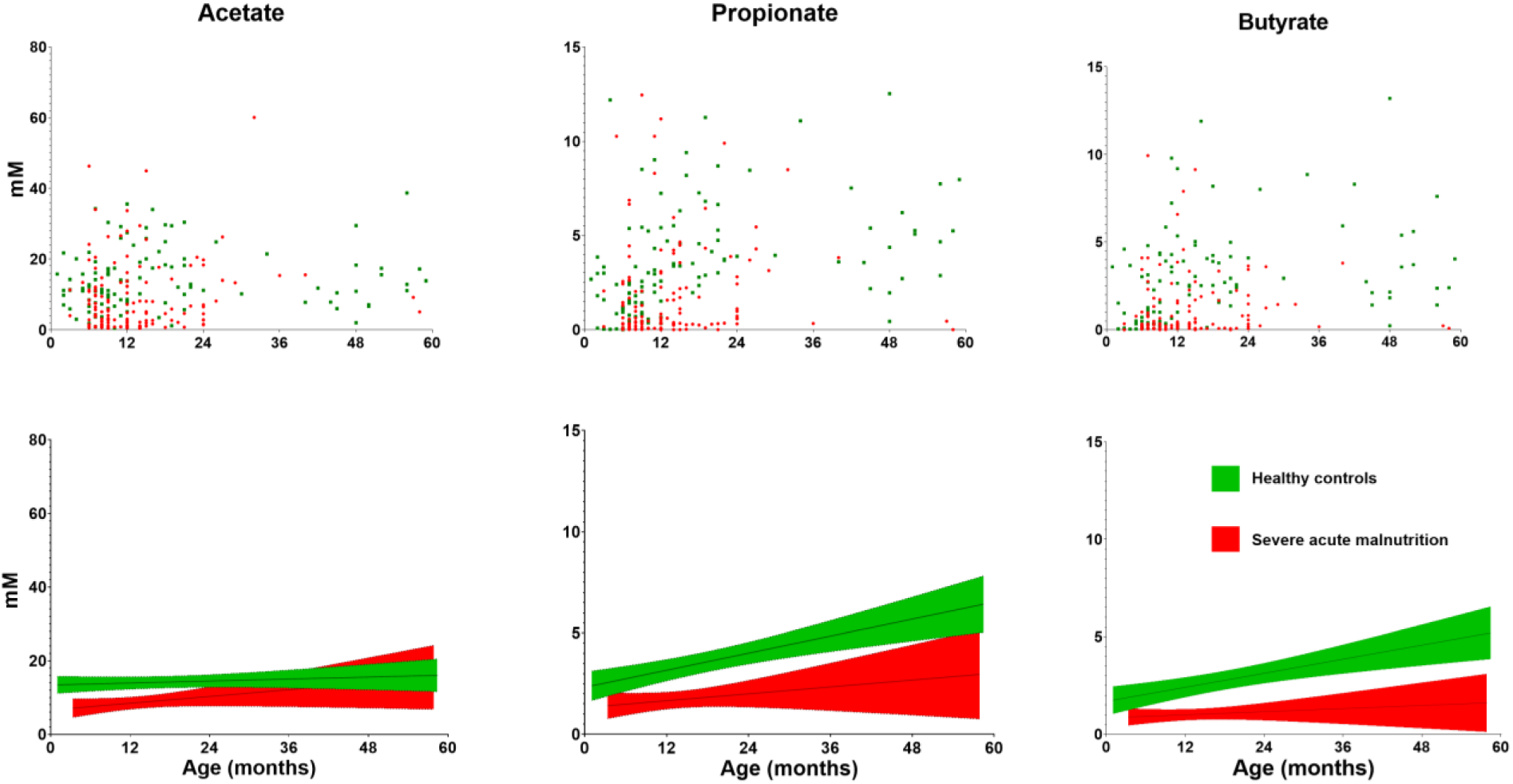
Variations in faecal acetate, propionate, and butyrate concentrations according to age and nutritional status. mM: millimolar, Confidence intervals at 95% of regression line are shown; green area reflects healthy control; red area reflects children with malnutrition.

#### 3.3.4. Depletion of faecal butyrate was associated with depletion of bacterial butyrate producers

Finally, we analysed the relative abundance of butyrate-producing species to assess its correlation with faecal butyrate depletion. We found a consistent and significant ten-fold decrease in butyrate-producing gut microbes in SAM compared to healthy controls (median, 2.1% in healthy controls versus 0.23% in children with SAM, *P* < .0001, Mann-Whitney test, Figure 7). In particular, this was also confirmed for *Faecalibacterium prausnitzii* (median, 0.026% vs 0%, *P* = .0003).

**Figure 7.**
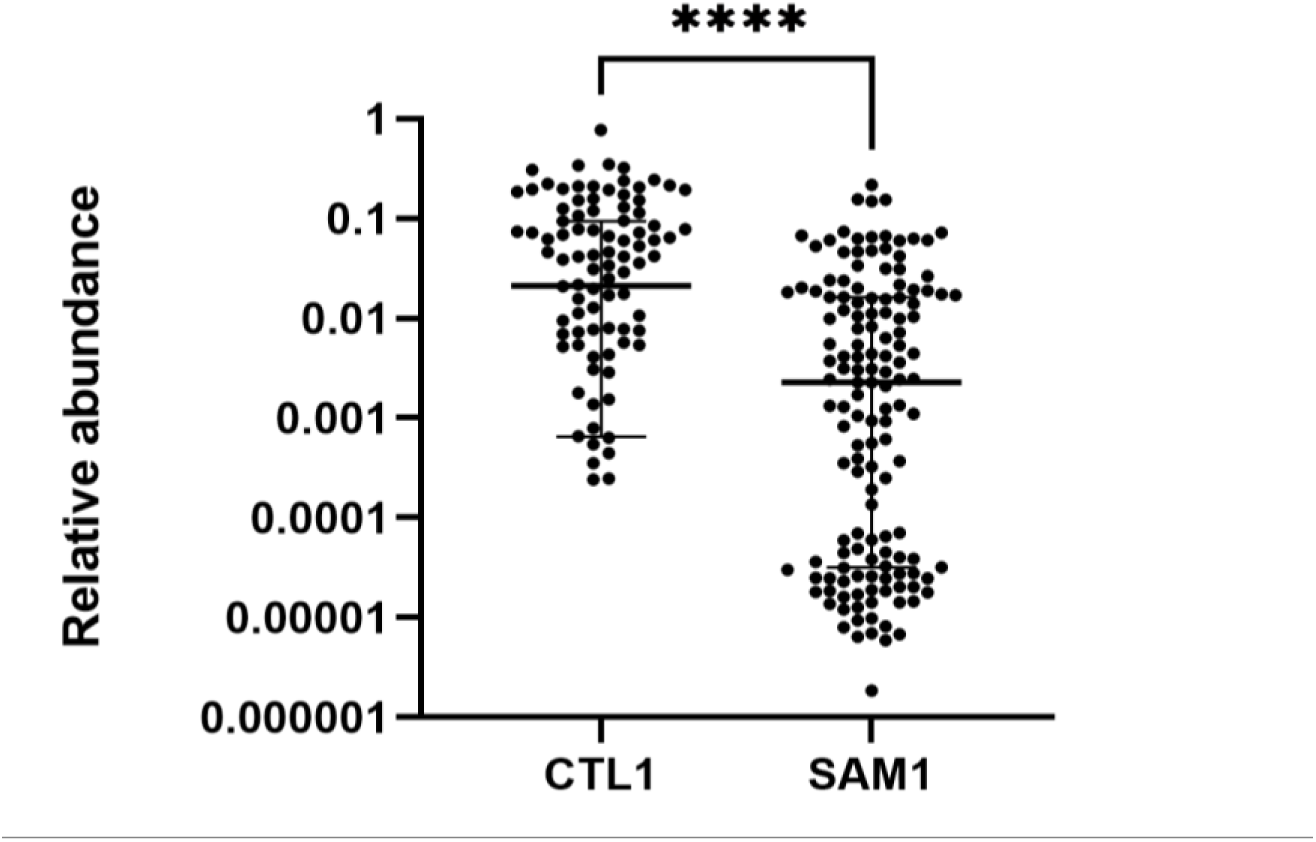
Relative abundance of butyrate-producing microbes in healthy versus severely acute malnourished children. CTL1: samples of controls, initial sample, SAM1: samples of children with SAM, initial sample. ****P < .00005, Mann-Whitney test.

#### 3.3.5. Therapeutic diet does not reverse faecal butyrate depletion

For 34 malnourished children, a second stool sample was collected at the time of discharge and analysed for short-chain fatty acid concentration. Faecal butyrate levels were not corrected at discharge in these children (Figure 8). For instance, focusing on butyrate, only 1/34 (3%) of faecal sample collected at the time of discharge was within the 95% confidence interval of the butyrate levels of healthy controls according to age.

**Figure 8.**
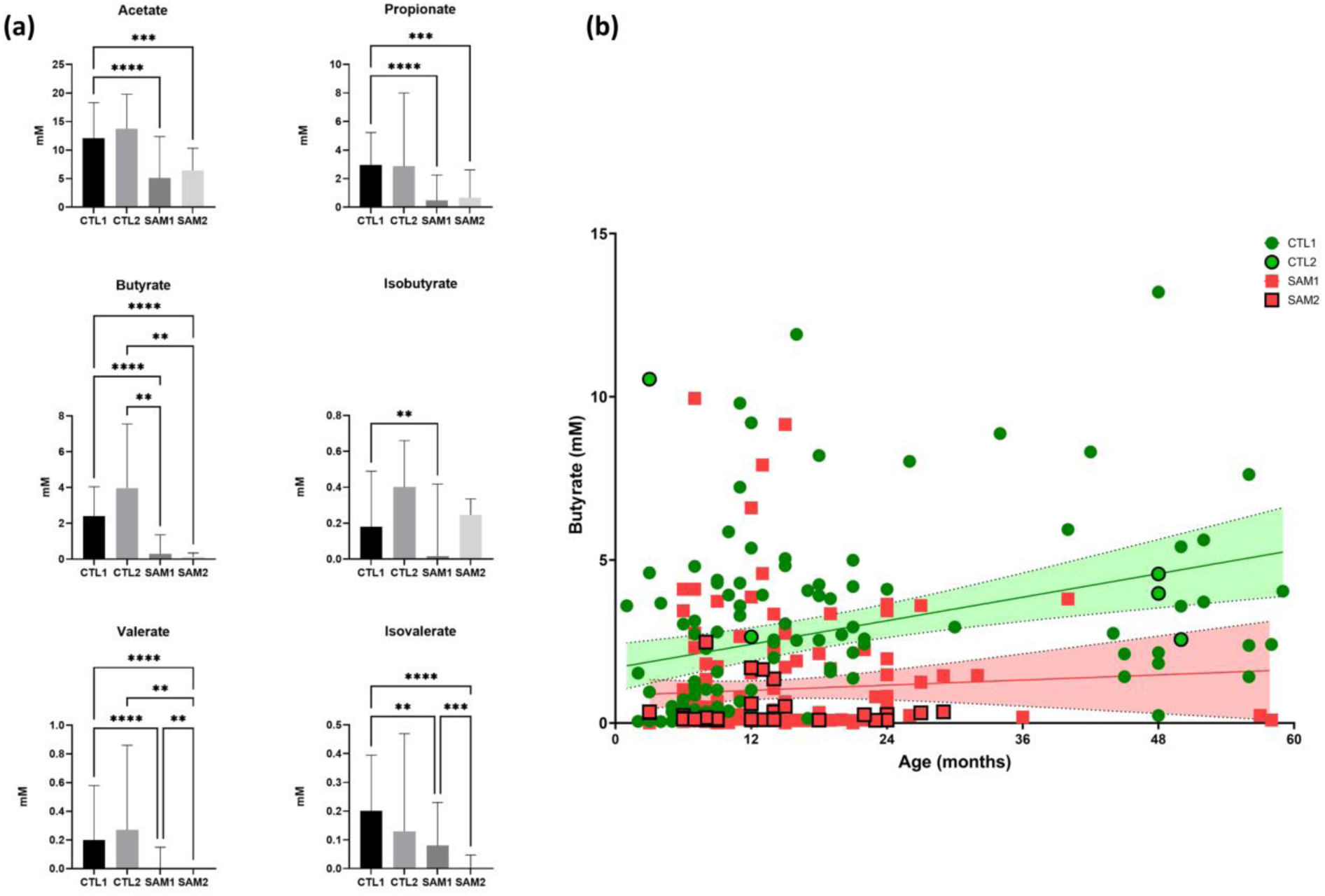
SCFA and Butyrate levels after therapeutic diet in the subgroup of children with SAM and healthy controls. CTL1: faecal sample from healthy controls at the time of admission, CTL2: second samples from controls (n= 5), SAM1: faecal sample from children with SAM collected at the time of admission, SAM2: faecal samples from children with SAM collected at the time of discharge (n = 34) (a) Comparison between the four groups. \*\**P* < .005, \*\*\**P* < .0005, \*\*\*\**P* < .00005. (b) Butyrate according to age. 95% confidence intervals of linear regression are shown. Arrow: only one sample from children with SAM at discharge is within the 95%CI of controls.

## 4. Discussion

We found that SAM was associated with a dramatic depletion of faecal SCFAs, particularly an approximately ten-fold decrease in faecal propionate, butyrate, isobutyrate, valerate and isovalerate. This was associated and consistent with a definitive confirmation of the relative depletion of gut aerointolerant microbes, and with a predominance of aerotolerant microbes in children with SAM, regardless of age. We identified samples of children with SAM with a complete loss of gut aerointolerant microbes. In such children, we were struck to find that a single potential pathogenic species, usually associated with sepsis and bacteraemia as *Escherichia coli* or *Morganella morganii*, may invade the gut microbiota, accounting for more than 75% of the total gut bacterial relative abundance. In another children, a single *Pseudomonas* species (*Pseudomonas lactis*) accounted for more than 90% ot the bacterial relative abundance.

*Pseudomonas* are strict aerobic bacteria and are usually not found in the gut. This suggests that the oxidative stress which has been reported for decades in SAM [5] is associated with a loss of the gut anoxic environment. Strikingly, all of these three species correspond to only two bacterial classes: Bacilli (for *Streptococcus*) and Gammaproteobacteria (for *E. coli* and *P. pseudoalcaligenes*), the two classes are associated with invasive infections in children with SAM in the literature [35,36]. This confirms the collapse of the aerointolerant microbiome and the relative massive enrichment in potentially pathogenic [37] aerotolerant and strict aerobic species in SAM. These findings are consistent with the relative proliferation of *Pseudomonadota* and especially enterobacteria in SAM, as previously reported in the gut microbiota [38]. We also confirmed the importance of potential pathogens, particularly *Klebsiella pneumoniae* [7,36,37] and *Fusobacterium nucleatum* [28,39,40]. While the effectiveness of antibiotics has been questioned in this context [41], these findings strengthen the inclusion of systematic antibiotics for SAM, consistently with our previous studies and meta-analyses [7,42]. As a febrile bacteraemia frequently reported in this context is being associated with increased mortality [35], systematic blood cultures should be undertaken. A recent meta-analysis by another team in Ethiopia confirmed the efficacy of amoxicillin in the initial care of children with SAM [43]. However, antibiotics which are more effective on enterobacteria, such as *Salmonella enterica*, *E. coli*, or *K. pneumoniae*, frequently resistant to amoxicillin (while *Fusobacterium* and *Streptococcus* are usually susceptible), should be tested in this context [7,35,36].

Anaerobic life was discovered by Louis Pasteur in 1861 [44,45]. The following year, he discovered butyric fermentation associated with vibrio, forming mobile spores capable of growing indefinitely in the absence of free oxygen [44,45]. In this context, he was the first to link butyric fermentation and anaerobiosis [44,45]. In humans, recent studies have confirmed that butyrate production relies on the gut aerointolerant *Lachnospiraceae* and *Ruminococcaceae* families and *Bacteroidetes* phylum [14]. Specifically, *Facealibacterium prausnitzii* and *Roseburia* spp. are important acetate utilisers and butyrate producers [46]. However, only a few studies from our centre have linked the depletion of gut aerointolerant microbes to SAM [4,7,15–17,25,37]. To our knowledge, this study is the first to link oxidative stress-driven dysbiosis, responsible for aerointolerant collapse, to the depletion of gut metabolites in a human disease (SAM).

Butyrate has been identified as a key microbial metabolite with several beneficial effects for mammal health, notably for regulation of liver fat and prevention of metabolic syndrome [10,11]. Butyrate was initially identified by French chemist Michel Eugène Chevreul in 1823 from fermented milk (“butyric” referring to ”from butter”) [47]. Butyrate has been recognised as a pleiotropic molecule in mammals with epigenetic properties (histone deacetylase (HDAC) inhibitor), binding to free fatty acid receptors (FFAR) 2 (GPR43) and 3 (GPR41), GPR109, and to peroxisome proliferator-activated receptors (PPARs) [11]. Butyrate activates peroxisome proliferator-activated receptor-gamma coactivor-1 alpha (PGC-1α), thus preventing high-fat diet adiposity and insulin resistance [48]. Dysfunction of PGC-1α is associated with mitochondrial dysfunction, fatty liver disease as observed in non-alcoholic steatohepatitis, and kwashiorkor [7]. Therefore, the protective effect of butyrate against oxidative and mitochondrial stress and the accumulation of fat has been repeatedly demonstrated [49],

specifically in the liver [50]. The critical and instrumental role of butyrate is further supported by the fact that Attia *et al*. [12] linked faecal butyrate depletion to mortality in children with SAM. Furthermore, in the present study, we found a positive correlation between butyrate and propionate levels and age in healthy children. This suggests that butyrate, associated with mortality in SAM [12] and propionate do represent the maturation of a healthy aerointolerant gut microbiota. This is not the case for acetate. The importance of butyrate for the normal growth and development of children in low-income countries is also demonstrated by the fact that Kort *et al*. [26] recently reported that the butyrate-producing gut bacterium *Coprococcus eutactus* was a predictor for language development in three-year-old rural Ugandan children.

In addition to butyrate depletion, we determined that gut valerate and isovalerate were also strongly affected by SAM. Valerate is produced from succinate and taurocholic acid, and contributes significantly to HDAC inhibition [51]. It is able to preserve gut barrier function [52] and inhibits *Clostridioides difficile* [53].

To our knowledge, this study is the first to systematically evaluate faecal SCFA diversity in children with SAM and controls. The role of SCFA as an energy source, in the development and maturation of intestinal cells, and the regulation of gut immunity [11,54] can explain digestive disorders and gut immune deficiency with a high risk of enteric sepsis in children with SAM [35,36]. Butyrate-producing microbes have complex requirements including several vitamins [55], frequently lacking in the diets of children with SAM.

We also demonstrated that aerointolerant gut prokaryotes are able to maintain butyrate production in the presence of oxygen in a medium supplemented with three critical antioxidants: ascorbate, uric acid, and glutathione [56]. This indicates the critical role of antioxidants in the gut environment, not only in the plasma and in human cells. Future studies could assess which antioxidants are the most important in the therapeutic diet to restore and prevent the loss of the healthy mature gut aerointolerant microbiota (HMAGM – [4]) and gut butyrate production. The association of low faecal butyrate and SAM mortality [12] strongly suggests that butyrate is at least a marker and possibly one of the key metabolites critical for the survival of children with SAM.

The depletion of aerointolerant microbes found in Malian children confirms all our previous studies on SAM using both culturomics and 16S amplicon sequencing in different countries (Senegal and Niger), and our comprehensive review of the literature [4,7,15–17]. However, the present study is much larger than previous published studies focusing on the aerotolerant/aerointolerant imbalance in SAM. The robust case-control design with successful age and sex group-matching enabled us to obtain unprecedentedly robust findings (Type I error < 10^-21^). We were surprised to find that age was not a confounding factor for the MAIPI, i.e. age did not impact aerotolerant/aerointolerant ratio in our study (Figure 2). This suggests that aerointolerant predominance is a key feature of a healthy gut microbiota early in life.

Here, we confirmed the collapse of the aerointolerant microbiome and oxidative stress in children with SAM, with a complete loss of aerointolerant microbes in some malnourished children. This alteration was associated with a collapse of short-chain fatty acids, whose role is increasingly recognised as crucial for children’s health and development by preserving mucosal barrier integrity, inhibiting pathogens, and contributing to immune and cognitive development. Our results confirm those of Attia *et al*. [12] and suggest that faecal butyrate could be used as a marker of dysbiosis severity and a predictor of mortality in children with SAM. In addition to the assessment of aerointolerant predominance by sequencing of the faecal microbiota, butyrate production capacity (butyrate kinase), and correction of the propionate and butyrate profile could be proposed for screening and follow-up of the treatment of these children. Failure to correct these markers with diet alone will make it possible to identify children in whom treatment including digestive microbes capable of producing propionate and butyrate could be proposed.

A link between deficient diet, antioxidant depletion [5,7], aerointolerant gut prokaryotes depletion, butyrate depletion, peroxisomal and mitochondrial dysfunction, and liver steatosis is thus suspected in children with SAM. This link, if demonstrated, would open avenues for SAM prevention and management. In the future, studies may investigate whether gut microbiota profiling may help guide antibiotic therapy in malnourished children with sepsis as, for instance, *Morganella morganii*, representing > 75% relative abundance in one child is naturally resistant to amoxicillin (AmpC β-Lactamase). After initial management with blood cultures and antibiotics if needed, future management of children with SAM should focus on restoring the aerointolerant microbiome and gut production of the key metabolites such as propionate, butyrate and valerate for children’s survival, and their metabolic and cognitive development [26]. Gut microbiota-targeted antibiotics for invasive species, and the restoration of aerointolerant microbiota and faecal butyrate could be part of future therapeutics in SAM.

## Author contributions

conceptualisation, OKD and MM; methodology, OKD and MM; software (MetaGX), AC, AL and MM; validation, MTA; formal analysis, AC, AC, AL, and MM; investigation, AC, SK, MTA, NA, AHT, SN, AKK, and IT; Resources, IT; data curation, AC, SK, and MM; writing—original draft preparation, AC; writing—review and editing, MTA, and MM; visualisation, AC and MM; supervision, SN, MAT and MM; project administration, OKD, and MM; funding acquisition, OKD and MM. All authors have read and agreed to the published version of the manuscript.

## Funding

This work was funded by ANR-15-CE36-0004-01 and by ANR “Investissements d’avenir”, Méditerranée Infection 10-IAHU-03 and was also supported by the Région Provence-Alpes-Côte d’Azur. This work received financial support from the Mediterranean Infection Foundation.

## Institutional review board statement

The study was conducted in accordance with the Declaration of Helsinki and was approved by the ethics committee of the Faculty of Medicine and Odonto-Stomatology of the University of Bamako, Mali, under No. 2014/46/CE/FMPOS on 22 May 2014.

## Informed consent statement

informed consent was obtained from all legal representatives (parents) involved in the study.

## Data availability statement

Data are available upon reasonable request.

## Conflicts of interest

the authors declare no conflict of interest.

## Acknowledgements

The authors thank Julien Paganini and the Xegen company (https://xe-gen.fr/).

## Notes

### Competing Interest Statement

The authors have declared no competing interest.

### Funding Statement

This work was funded by ANR-15-CE36-0004-01 and by ANR Investissements d avenir, Mediterranee Infection 10-IAHU-03 and was also supported by the Region Pro-vence-Alpes-Cote d Azur. This work received financial support from the Mediterranean Infection Foundation.

### Author Declarations

Ethics Committee of the University of Bamako Faculty of Medicine and Odon-to-Stomatology waived ethical approval for this work on 22 May 2014 (2014/46/CE/FMPOS). Informed and signed consent was obtained from all legal representatives (parents).

